# Infection Prevention and Control practices; an exploratory qualitative study of experiences of health care providers and support staff at the University Teaching Hospital, Lusaka, Zambia

**DOI:** 10.1101/2022.03.23.22269760

**Authors:** Martin Magule Zimba, Chanda O. Dorothy, Chama Mulubwa, Alice Ngoma-Hazemba

## Abstract

**Background:** Infection Prevention and Control is a public health concern to abate Hospital Acquired Infections. Hospital Acquired Infections are known to complicate clinical care, increase length of stay in hospitals and particularly have disturbing effects on patients’ recovery as well as devastating effects on health care facilities, especially in low resource settings. However, with effective infection prevention practices, the Microorganisms can be eradicated through proper hand hygiene practices of the health workers and proper waste management and disposal/adequate environmental sanitation.

We explored the experiences of health care providers and support staffing infection prevention practices in selected clinical areas at the University Teaching Hospital, in Lusaka.

**Material and Methods:** This was a qualitative study that used an exploratory research design. The exploratory design allowed the participants to give in-depth narratives from their work environments as well as enabled the researcher to observe their Infection Prevention Practices. The observations provided an opportunity to make varied conclusions on compliance to infection prevention practices. The researcher collected data using an interview guide from the thirty (30) study respondents. Observations were conducted in their work environments to strengthen the conclusions made. Audio recordings were transcribed verbatim and then imported into Nvivo Version 11 for data management and analysis. Thematic analysis guided the conclusions made in the study.

**Results:** Three main themes emerged from the data analyzed: These were existing guidelines on selected Infection Prevention and Control guidelines; Knowledge and use of the Infection Prevention and Control guidelines, and compliance to Infection Prevention and Control guidelines by health care providers and support staff. The participants demonstrated good understanding of the benefits of observing standard IPC guidelines in the workplace, barriers to compliance were reported in almost all the sites. Staff turnover, limitations in infrastructural, and heavy clinical workloads led to lapses in compliance with Infection Prevention Practices.

**Conclusion:** The study recommends the setting up of an effective and efficient Infection Prevention and Control Committee at the University Teaching Hospital. The committee should oversee the proper functioning of all safety and occupational Health activities and compliance to all infection prevention and control guidelines in the clinical areas and hospital environment by ensuring effective hand hygiene practices using elbow–operated taps and proper medical waste disposal starting with the use of foot-operated bins and compliance to all existing infection prevention guidelines in all clinical areas within the hospital..

## BACKGROUND

Hospital Acquired Infections (HAIs) can be prevented by adequate and less sophisticated surveillance and control measures [1]. Hospital Acquired Infections (HAIs) are known to complicate clinical care, increase length of stay in hospital and particularly have disturbing effects on patients’ recovery as well as devastating effects on health care facilities, especially in resources limited settings [2-5], Therefore, Infection Prevention and Control Guidelines (IPCGs) are essential in the delivery of modern health care services to reduce HAIs. Effective implementation of IPCGs is crucial to reducing the transmission of HAIs; however, there is limited information on comprehensive assessment on why health care providers fail to adherence to IPCGs in some settings especially the resource-limited countries. In 2010, the World Health Organization (WHO) reported that only 23/147 developing countries have a functioning surveillance system for HAIs, which is a core part of infection control programs [6], thus stressing the importance of Infection Prevention and Control (IPC) committees in health institutions. There are a number of international pronouncements [7] to support low and middle income countries to build and implement IPC initiatives in their health care settings. Some of the initiatives include among others; hand hygiene, sterilization and disinfection of medical material, aseptic techniques, prevention and management of injuries from sharp instruments, early detection of disease and isolation precautions such as patient placement, use of personal protective equipment, and waste management. Despite the efforts to address this gap, HAIs, in health care settings remain a significant threat to the quality of patient care.

Alp E, and others [8] established that the risk of HAIs is greater in low-middle income countries whose magnitude remains underestimated or even unknown largely because HAI diagnosis is complex and surveillance activities to guide interventions require expertise and resources. The university teaching hospital (UTH) in Zambia is the national referral hospital at the tertiary level of care. It’s a large structure spread over one and half kilometers stretch with approximately over 3, 000 Medical Personnel and about a bed capacity of about 2, 400. Studies conducted at the university teaching hospital on IPC for example, demonstrated that up to 37 percent of nosocomial infections are directly attributable to cross transmission of resistant organisms across patients and health care providers in this facility, further indicating that health workers at the UTH generally have high microbial contaminations [9].

This research sought to understand health care providers’ and support staff’s experiences on infection prevention and control practices and compliance with recommended IPC procedures at this tertiary health institution.

## METHODS

### Design

This was an exploratory qualitative study explore experiences with IPC practice and participants were drawn and interviewed from their work environment which allowed them to give an in-depth narration of their work spaces and enabled the researchers to observe activities as well. The observations provided an opportunity to make varied conclusions on compliance with IPC guidelines.

### Study population

Study participant included management, nurses, midwives, doctors and support staff from the selected sites of the University Teaching Hospital.

### Recruitment of participants

A purposive sampling method was used to recruit all participants from the Adult and the Women & Newborn hospitals. The participants were recruited because of the nature of the work they performed on daily basis. In addition, the managers were recruited to provide guidance on how they managed the IPC policy guidelines for their institution.

### Data collection methods

Data were collected through in-depth interviews conducted in June 2017 among 30 health care providers and support staff from the selected sites at the University Teaching Hospital, in Lusaka, Zambia. Oral and written informed consent were obtained from each participant after explaining the purpose, benefits of the study and participants were assured of no risks, and how the information would be used to improve infection prevention practices. A semi-structured interview guide was used to moderate the discussions.

### Data management

All interviews were audio recorded and transcribed verbatim. A saturation point was reached when no new information or themes were coming out from the interviews.

### Data analysis

All transcripts were entered into Nvivo-10 software. Thematic analysis guided the conclusions made in this study. The principal researcher proceeded by familiarizing himself with the data and emerging major themes were used to identify supporting themes for in-depth descriptions on experiences after a series of reading and rereading the transcribed data. The coding was achieved through indexing the presence of each theme and selecting direct quotations to support subthemes. There after data triangulation was done by cross-referencing between categories of participants in the study.

### Ethical considerations

Permission and ethical approval were granted by National Health Research Authority and the Biomedical Research Ethics Committee **(*REF. No. 059-06-17*)**. Written informed consent was obtained from every participant and they were reassured of confidentiality, however, were free to withdraw from the study without any penalties.

## RESULTS

A total of 30 health care providers and support staff including 2 administrative staff at hospital level participated in the study. Participants were categorized according to their current designation. Participants were derived from health professionals, including health support staff at the University Teaching Hospital; 10 Doctors, 11 Nurses, and 7 Support-staff participated in the study. 2 Administrative staff was later included to clarify certain claims by the participants. The fundamental themes emerged from the scripts of the 30 participants in the research study.

**Table 1.** Characteristics of participants in interviews regarding their perceptions and experience on infection prevention and control in UTH, 2018 (N□= □ 30)

| Part. Code | Gender | Number of Part. | Experience. Yrs. Served | Clinical area Serving | Clinical Area Experience | QUALIFICATION |
| --- | --- | --- | --- | --- | --- | --- |
| IDI. Doctor | Males | 05 | 5 Yrs. above | 7 Yrs. IMCU | Med-Surgical | DEGREE |
|  | Females | 04 | 5 Yrs. above | 3Yrs. above |  | MMed |
| IDI. Nurse | Males | 04 | 5 Yrs. above | 5 Yrs. NICU | Pediatrics | RNs |
|  | Females | 08 | 8 Yrs. above | 3 Yrs. Above |  | RNs |
| IDI. Maid | Females | 04 | 18 Yrs. | 10 MTN | Maternity | GCE |
| IDI. Porter | Males | 03 | 5 Yrs. above | 7Yrs THTR | Theatre | GCE CRAFT |
| ADM |  | 02 | 5 years. above | 6 Yrs. Mgt. | None | DEGREE |
| **TOTAL** |  | **30** |  |  |  |  |

**Note.** RN = Registered nurse, MTN= Maternity, ICU = Intensive care unit, IMCU = Intermediate care unit, NICU = Neonatal Intensive care unit, Mgt. = Management

## EXISTING GUIDELINES ON SELECTED INFECTION PREVENTION AND CONTROL

For this study, the guidelines were described in the context of hand washing, waste management and disposal as well as the workstations in relation to cleanliness, bed spacing and isolation of patients with infectious diseases. The following sub-themes describe participant awareness of existing these guidelines in their respective work environments:

### Hand Washing

Regarding availability of IPC guidelines on hand washing, the Health Care Providers were able to explain that cleaning heavily contaminated hands with an antiseptic before patient contact reduced HAI transmission of contagious pathogens. However, they were quick to highlight the limitations due to lack of running water in their workstations.

> *Washing of hands in medical practice is important as it obstructs the spread of infectious bacteria in the hospital environment. This is achieved by the washing of hands with soap and clean running water to rid of all dirty and germs from the hands before handling anything else. (IDI Maternity Nurse 15)*
>
> *…Many diseases and conditions are spread by not washing hands with soap and clean, running water. If clean, running water is not easily accessible as is common here in the surgical adult wards; cross contamination and HAIs thus have a bearing of the recovery of the patients admitted to these wards. (IDI Ward Manager Surgical Nurse 01)*
>
> *…we most of the times have no access to constant supply of running water; either the taps are broken or leaking and were forces to shut to avoid floods and so we store water in containers. Yet despite doing so, we risk contamination of the same stored water when one fetches using a cup to wash hands. Running water sources and sinks are most of the times broken because they are overwhelmed*…*so we provide water sources in terms of bucket taps for our staff but it’s not enough. (IDI Ward Manager-Surgical Nurse 01)*

Despite the above context related to the importance of hand washing, the participants demonstrated that there is an indirect contact route of transmission from one patient to another. This was emphasized by the fact there is lack of infrastructural compatibility, implying that the right facilities (hand wash basins) needed to be installed if hand washing was to be done correctly. A health care provider from a neonatal ward explained: *“The water points are far and those close to our working areas are broken most of the times due to use overloads…hand basins are broken we mostly depended on the bucket-taps as wash basins which is an indirect contact route for pathogens in clinical areas. Normally we should have had elbow or wrist lever operated mixer or automated controls not theses “ordinary taps” where we touch even after washing of hands rendering them heavily unsterile. This happens in that after washing hands, the user gets them dirt in the process of closing the tap. (IDI Neonatal Female Nurse 05)* It was also noted in the a Surgical ward that some health care providers fail to wash their hands due to workload and constant inflow of patients in these wards.

> *…due to a lot of work we sometimes forget to wash our hands as we concentrate on saving the lives of many critically ill patients in our department. Regardless of availability of soap, most staffs do not use it since we use glove. (IDI Surgical Male Nurse 02)*

### Medical Waste Management (MWM) and Disposal

Participants acknowledged that Medical Waste Management was considered as one of the important aspects of infection prevention & control. As dictated in the guidelines, they were of the opinion that segregation of waste at source is the best practice, yet this was not the case. Those doing the sorting (of mixed waste in large quantities) especially the cleaning staff, expressed concern and claimed that there was negligence by the professionals in managing medical waste. Poor handling and disposal of medical waste highlighted a lack of knowledge among the participants. A cleaner in a Maternity ward explained how the waste they manage is not always segregated:

> *We have no separate storage points for biohazard from the general domestic waste. …We dumped in that (big Black) container, which is daily, emptied by us for disposal. Nonetheless, used syringes, blood drip sets, medicines bottles and urine bags are sometimes found inside this general waste container…Hence it is better to practice the process of segregation at point source. (IDI Maternity Porter 20)*

The ward managers on the other hand explained that they failed to follow the waste segregation protocols because of lack of reminders on protocols on posters to direct safe waste disposal in designated places. It was observed that in some places, there was lack of awareness among the staff in management positions like doctors and senior administrators to emphasis clarity about recommended practices especially regarding segregation of waste at the point of generation. Some remarks presented here indicate their experiences with waste management.

> *…we sometimes fail to comply with infection control guidelines, instructions or protocols and hence the learners graduate with poor knowledge. As you’re aware this is a teaching hospital, therefore there’s need for constant reminders that waste management is part of good practice (IPC) to mitigate spread of HAIs. (IDI Neonatal Doctor 09)*

Further, medical waste sorting and disposing in color-coded/labeled containers was a challenge and it was observed that there was a mixture of medical and domestic waste in the places visited. Despite having written notices indicating the treatment of waste and disposal, HCP still disposed medical waste without sorting. A nurse in a surgical ward explained: *We dispose all the medical waste in a single bin except the sharp objects. Soiled things and filth are disposed in the black bucket. Water [liquid] is kept in the red bucket…I don’t know much about the codes for each ward…to tell you the truth Sir…as health care providers our focus is on diagnostics and treatment of patient …so we pay less attention to the sorting of medical waste and proper management when we have maids to clean and potters for that job …instead we pay attention to treat patients. (IDI Surgical Ward Nurse 26)*

A medical doctor from a neonatal ward added: *“What happens when the waste is not covered the same goes in the same lift. The doctors are going in the same lift, patients are going in the same lift and they are taking it (waste) open. When we go on rounds at nine A.M. that waste transport man comes with his cart and the foul smell and it is open, all the pads and what not may be the source of infection also”. (IDI Neonatal Doctor 09)*

Lack of proper waste management poses a risk for those who manage the waste in the settings. A cleaner lamented: “*As women in the cleaning department, we are responsible for removal of the waste; we usually are at risk of contracting disease…we lack PPE on our side which makes us vulnerable to infections. Even the health professionals often don’t consider us important partners who can be worthy considering in day-to-day duties but considered low lives. (IDI Maternity Maid 13)*

### Health Care Settings and Bed Spacing

Participants reported that the health care settings and workspaces were a source of infectious agents such as the virus, bacteria, or other microbes. Coupled with human traffic of patients and their visitors, safety was compromised because everyone uses static and moveable infrastructure:

*… People are one source of germs including: Patients, Healthcare workers, Visitors and household members. …infrastructure I mean that there is poor environmental cleaning of e.g*., *bed rails, medical equipment, countertops, and tables. … You find that there are wet surfaces, moist environments, and biofilms, faucets and sinks, and equipment such as ventilators. Indwelling medical devices (e.g*., *catheters and IV lines). Dust or decaying debris (e.g*., *construction dust or wet materials from water leaks). (IDI 19. Surgical Ward Nurse)*

In some settings, a lack of isolation rooms where burn wound patients should be housed for example are poorly managed. Wound cleaning procedures were not entirely applied because of inadequate spacing, overcrowding and poor ventilation systems. Participants were concerned about the lack of space and ventilation systems and about overcrowding in the key areas of risk where patients needed enough space. The following narratives were by staff from surgical wards:

> *We do have an isolation room in the wards but the ventilation system is not really good. There is not enough space at all for all special cases in this areas, so separation is not done accordingly.…the facility is not a big enough facility to separate Open wound patients from other patients, so all patients are admitted to the same ward. (IDI 6 Surgical ward Doctor)*
>
> *…We don’t normally separate patients because we give preference to those that come earlier to take the front seat. If we separate patients with open wounds, it may discourage other patients that came early from theatre like we are giving special preferences. (IDI 6. Surgical ward Nurse)*.
>
> *…There is are no air is extractor from the room, and we experience frequent shortage of basic alcohol saline and other wound cleaning supplies. (IDI.09 Surgical Wad Doctor)*

## KNOWLEDGE AND USE OF IPC POLICY GUIDELINES

To explore how the health care workers applied the knowledge of IPC guidelines, participants in this study were asked about what they knew and how the used the information to prevent HAI. This knowledge is described in the context awareness of IPC,

Participants demonstrated lack of standard precautions knowledge in waste segregation and sub-optimal use of personal protective equipment.

> *We honestly have poor Knowledge on Infection Prevention Control. Most of us have not undergone IPC training. …We need in-service courses to remind us of the correct procedures. We lack knowledge on how to control infection and without the support of complementing infrastructure and adequate supply of consumables such as soap, were far from winning against HAIs but were trying. (IDI Maternity Nurse 15)*.

However other participants had some average knowledge levels but explained with reasons why they were non-compliant citing increased workload and poor risk perceptions and practice.

> *We mostly know what is supposed to be done but its bad attitude at play, fatigue and lack of motivation…coupled with bad working environment that is not support IPC procedures…we’re just put off sometimes*. (*IDI Surgical Female Nurse 03)*.

The in charge re-echoed this and explained that, there is lack of refresher courses on IPC and personnel with no sanctions by authorities for noncompliance does not command a sense of responsibility in many heath care providers and therefore such can only be enforced by policy formulation and implementation through in-service training workshops. Ensure also curriculum development by colleges and universities to highlight the importance of preventive medicine.

> *Most of us health workers are well trained; we need to have refresher courses and sometimes it is just bad attitude towards work. But most students are ignorant because there mentorship during training and we lack administrative strict adherence policy to this effect. Students who start training at the hospital don’t know what the color codes mean unless their supervisors explain them. There is a risk of infection to patients and themselves when students enter a zone without knowing how to dispose of medical waste. (IDI Neonatal Female Nurse 06)*.

The current health financing system is doggy when it comes to funding programs against the burden of HAIs. Two ward managers highlighted the importance of building financial mechanisms at UTH to influence infection prevention programs to take center-stage and avoid antibiotic remedial measure to HAI.

> *…Currently MoH officials, mainly those who have a non-medical background tend to cut resources planned for IPC activities because they lack knowledge on IPC. …You find MOH prioritizing antibiotics and other medicines as remedial interventions to such infections instead of dealing with the root cause and strengthen IPC. Thus, infection control receives a low priority IPC. We hope to see a change in effort towards preventive medicine with the establishment of public health division at Ndeke. (Ward Manager.Nurse.01)*

Policies for eliminating Hospital Acquired Infections and Policy directive outlining practices are required in order to minimize the risk of patients, visitors, students, health care providers and support staff working in the hospital environment against communicable diseases. The primary purpose of the policy is to provide leadership, manage and control hospital acquired infections (HAIs), and minimize the adverse health impacts and reduce its burden. Participants did not know of clearly outlined IPC policy that can be enforced. Therefore no officer has been made answerable for any wrong doing in terms of HAIs.

> *When cases of negligence are reported …it comes out just as rumor… people say that big hospitals don’t report their cases in order to avoid trouble. We lack policy for ensuring patient safety when such cases come up. Usually management is quick to defend the health worker who abrogates these IPC rules to protect the image of the institution. (IDI Maternity Doctor 21)*

The study revealed inadequate education and training as an influencing factor and this evidenced by the following direct quotation from the participants:

> *…most of us never get training on infection prevention and control issues… we have inadequate knowledge because in service training is mean for New-Comers (recruited personnel). We sometimes do not know what to do and just act according to our conscious. (IDI Maternity maid 19)*
>
> *…we can’t claim we have the best knowledge…our experience has been that such training programs are meant for those in professional positions not us… maybe were not as important. (IDI Theatre Maternity Porter 20)*

## COMPLIANCE WITH IPC GUIDELINES

Compliance with IPC policy guidelines was described in the context of administrative controls on IPC, roles and responsibilities of IPC committee and attitudes of HCPs towards application of IPC guidelines. A more comprehensive description with participant quotes is provided in the following subthemes:

### Administrative Controls on Infection Prevention and Control

Participants believe that lack of strict control and penalization as a response to reporting cases of Hospital Acquired Infections has led to non-reporting of infections and consequently no data. According to the participants, the Ministry of Health has no Officer in-charge of Infection Prevention & Control; therefore, IPC issues are resolved independently in different hospital divisions (wards). They also explained that “If there is no internal person who brings issues to the attention of the hospital management, then problems remained unsolved” and therefore infection control remains neglected.

> *IPC plans are not well coordinated without administration taking a lead. MoH lacks leadership in terms of controlling and allocation of resources to fight HAI; I feel they should create a new position which will be in-charge for coordinating infection control programs and plan at the national level. Nothing will ever materialize if Ndeke (MoH Head Quarters) has no such Coordinator. (IDI Surgical Doctor 24)*

Although participants acknowledge to a number of means of transmission, much was eluded to the fact that there is no proper bed spacing and this does not support the strict adherence and so management of HAIs and they can only do as much.

> *…I feel the reasons for noncompliance may be due to the poor infrastructure in the wards. As you can see other patients are on the floor, and as result we have situational heavy workload, overcrowding, complexity of care, and of course lack or unreachability of hand hygiene resources like washing basins which cannot be over emphasized on their dilapidated nature. (IDI Surgical Doctor 24)*

### Roles and Responsibilities of the Infection Prevention & Control Committee

Study participants explained that there is no IPC and Control of Hospital Acquired Infections. They claimed that this was the biggest challenge to enforcing standard IPC precautions. Without an office to report such cases to, it is impossible to even follow an offender of these guidelines. Positive leadership is considered a prerequisite in effectively controlling infection.

> *MoH has no staff in-charge of HAI control policy and, therefore, infection prevention control issues related for instance to HIV, blood transfusion, sterilization of equipment, etc. are solved independently in different wards. If there is no internal person from the wards who brings issues to the attention of authorities, then problems remain unsolved, and that’s how infection prevention control has remained neglected. (IDI Surgical Nurse 28)*

Participants appreciates that the standard protocols are in place for health care providers but told this researcher that there is no one to enforce them. They said that there is need to formulate policy at national level that would cut-across all the hospital in order to mitigate HAIs. Most of them recalled of past practical efforts by the Central Board of Health to form IPC committees mandated to enforce and conduct inspection to this effect. Over the years this effort has failed to materialize because there is none responsible to spearhead this cause at administration level and now is just on paper.

> *We do not have an active, stand-alone, national IPC programs which could define clearly the objectives, functions and activities for the purpose of preventing HAI and combating HAI through IPC good practices. Until this is put in place, I feel that these research findings and talks of HAI & IPC will remain just as an academic exercise. (IDI Theatre Maternity Nurse 16)*

This researcher could not find information or records of incidences nor validate of its existence as informed that man who was in charge had retired. Surveillance data therefore is needed to guide the development and implementation of effective interventions controls if serious decisions have to be made on reducing HAIs. Without management taking a lead, it will remain difficult to fund activities to enhance IPC compliance.

> *I remember in 1986, UTH management was concerned with nosocomial infections hence it set up a hospital infection control committee HICC…we saw everyone minding and doing the right thing. It later changed to Safety Occupational and infection Prevention Committee (SOIP) with an objective of monitoring incidence and prevalence of HAIs among the workforce and visitors. (IDI Theatre Maternity Doctor 29)*

### Attitudes towards the application of IPC among the participants

A significant proportion of HCP working in wards felt that there is need for management to deploy more nurses even so with the introduction of four separate hospitals in order to facilitate provision of safe and quality hospital care. A few health care providers explained that guidelines are available in the departments, yet the attitudes of the health care providers are bad. Other participants explained that the IPC policy is just on paper; non much has been done to enforce the strict adherence of these guidelines.

> *…Guidelines are there, but some people are not implementing them in practice maybe because there’s no one to prompt us. We have not seen the person in charge who is IPC designated to handle any issues of hospital acquired infections. (IDI Maternity Doctor 21)*
>
> *…increased workload prevents me from doing the right practices, for example there are times when I should assist two or three women who are all in second stage of labour, I have to do things really fast even though I know that chances of me contaminating are high but were left with no better choice than hurry. (IDI maternity nurse 10)*
>
> *…most of the time, the management comes only when there is bad situations in the wards in relation to infection prevention and control issues. I think management believes things are fine down here and while things are not fine in relation to IPC measures. (IDI Theatre Nurse 09)*

The participants further explained that there is no reporting system on HAIs and no records were found by the researcher to this effect. They explained that there could be pieces of legislative guidance to avoid HAIs but that there is no enforcement hence, it is left to individual’s conscious acts when handling patients.

> *Reporting of HAI policy is not really there; we work as careful as possible and handle things as they come. I have not seen any anyone being punished for the wrong procedure in IPC…the ward managers is themselves not doing anything about the offenders of the IPC standards precaution. (IDI Neonatal Female Nurse 06)*

Other participants explained however that the IPC policy is there but failure to actualize it because there has been not much done to enforce it. No one has ever been punished for wrong doing. Emphasized that they have not seen the person who is IPC designated to handle any issues of hospital acquired infections.

> *…like there is a penalty for drivers who drive recklessly, in the same way there should be laws to punish those who do not adhere to guidelines and principles aimed at preventing HAIs, e.g. hand washing…strict rules are needed and forceful implementation should be there …from top to bottom if they are not following the rules, and then enforce the rules. (IDI Neonatal Doctor 10)*

## CHAPTER FIVE

## DISCUSSION

This study highlights that compliance with existing infection prevention and control guidelines is suboptimal among health care providers and support staff in the selected clinical areas of the University Teaching Hospital in Zambia. Overall, participants were aware of IPC guidelines, however, their lack of application of the guidelines at administration and IPC committee levels had a bearing on compliance with the protocols in the work environment. The findings are described in the context of the existing literature to provide a platform for opportunities to strengthen the health care system in mitigating HAIs in this and similar settings.

### EXISTING GUIDELINES ON SELECTED INFECTION PREVENTION AND CONTROL

Application of the IPC guidelines developed by the Centre for Disease Control for reducing HAI form the basis for preventing transmission of micro-organisms within a hospital facility [1, 10]. Although the guidelines were not on site, participants for this study reported that they heard about IPC guidelines during their professional training and were familiar with them. However, they were not aware of the existence of an Infection Prevention Control Committee in their institution. The role of IPC committee was to monitor and evaluate the IPC practices to inform management and the health care system. It was noted in this study that the practices IPC among the health care providers and support staff were not entirely independent of policy guidelines. For instance, the suggestion to customize policies required reasonable practical issues while taking account the differences under which HCP operated was evidence that the participants consider this part of the health care delivery system to be important. Some of the important guidelines that required serious considerations were hand washing, medical waste management and disposal as well as environment/health care settings/bed spacing.

Studies have shown that hands are the most important and frequent mode of transmission of infection in the health care setting because organisms are transferred through direct contact between an infected patient and a predisposed health care worker or another person. In the settings studied, there was a lack of infrastructural facilities to support implementation of this important aspect in IPC practice. These limitations have been reported in similar settings of the low-middle-resource settings and recommendations to step up the infrastructure have been made [4, 11].

Other scholars have reported some known and tested technologies in hand hygiene which remains to be the simplest and the primary measure to prevent HAI and reduce spread of multidrug resistant organisms such as Tuberculosis [12] and yet without appropriate facilities such interventions are ineffective in resource limited settings. Samuel & others reiterated that availability of IPC logistics such as running water, soaps and funds have a bearing on motivation and compliance with hand hygiene in preventing the spread of HAIs [13].

On the other hand, the guidelines relating to Medical Waste Management in Clinical areas was considered as one of the important aspects of infection prevention & control in the studied sites. For instance, there was a suggestion that segregation of waste at source is the best practice although not adequately implemented. There was a concern that the workers (especially the cleaners) who sorted the waste and felt that clinical staff were negligent in implementing IPC practices which could even predispose handlers to injury and spread of infection. Failure to follow the waste segregation protocols was attributed to lack of reminders despite posters reflecting safe waste disposal guidelines being present. Inadequate staffing in the selected clinical areas led to failure to comply to IPC guidelines on waste segregation, a feature reported in similar setting, an imbalance in clinical settings where the wards with the greatest need have fewer staff such as the maternity and delivery wards [4].

In addition, this study found that the environment in which the IPC guidelines were implemented was not limited to hand hygiene and waste management practices but also to settings and bed Spacing. This was based on the understanding that the environment is generally a source of infection in which the viruses, bacteria, or other microbes are found. This was understood in the context of lack of facilities (isolation rooms) where burn-wounds patients for example were supposed to be managed. Wound cleaning procedures were not entirely applied because of inadequate spacing, overcrowding and poor ventilation systems.

It should be acknowledgement that the University Teaching Hospital is an institution where clinical practice mentorship is taught for medical students and other clinical staff. In this study however, the authors observed that the theatre staff demonstrated good standard practice such as sterile fields where the Nurse in-charge produced a checklist and protocols that were well coordinated to avoid cross contamination, Benchmark for good IPC practice for the whole institution especially for training and mentorship Stone, (2013:1).

Using their infection control training, nurses play a vital role in creating a culture of patient safety. On the other hand, overcrowding of patients and limited space was reported and compromised good IPC practices and risked cross infection among patients [14]. In the settings studied, there was a high volume of patients and sometimes health care providers are overwhelmed which required additional measures to mitigate the risks of spreading HAIs.

### 5.2. KNOWLEDGE AND USE OF IPC GUIDELINES WITHIN THE WORKSPACE

Some HCPs exhibited sub-optimal IPC practices and there were lapses observed in the completion of certain protocols. Participants were quick to mention that the infrastructure was not supportive for IPC implementation. Concerning the source of IPC information, participants mentioned training schools while others learnt through departmental meetings, and hence barriers and challenges hindered their compliance to effective infection prevention and control. The key to narrowing the knowledge gap and sustaining good IPC practice in health care lies in the strengthening of national capacity to ensure the availability of relevant and high-quality health information and evidence to inform decision-making to formulate site specific workplace protocols [15]. Participants attributed the lapse to lack of proper coordination of IPC practice to the absence of leadership and non-availability of an active IPC Committee whose responsibilities among others is to formulate and implement IPC policy and other clinical procedural guidelines.

In recognition of the limitations, participants recommended the provision of in-service training for effective and efficiency infection prevention and control protocols in clinical practice. A medical consultant suggested that it would be essential to allow senior staff to inculcate a positive mindset towards controlling HAI in health care providers learners because UTH is a learning and teaching hospital.

### 5.3. COMPLIANCE OF IPC GUIDELINES

This study highlights that administrative controls, non-performing Infection Prevention and Control Committees, and application of IPC practices provide the context within which the participants experienced a lack of compliance with IPC. Regarding management controls, participants explained that inadequate finance was among factors affecting IPC compliance [16]. This is in agreement with other researcher who stated that Africa carried 25 percent of the world’s disease burden, yet had only 3 percent of the world’s HCP and 1 percent accounting for nothing of the world’s economic resources to meet IPC challenges [17]. Resource constraints have thus facilitated in cutting corners leading to sub-optimal practices with consequences for HAI prevention [1]. The findings of this study suggest that when allocating resources, policymakers choose other vital clinical demands such as drugs, and other laboratory consumable supplies over infection prevention and control expenditures. This is on the basis that IPC activity spending is often misconstrued as being a waste of resources. In this instance, other studies have recommended exploring medical staff’s experiences, attitudes and practices towards IPC practices in order to address these limitations in decision and subsequently prevent nosocomial infections in this and similar settings [18]. A lack of valid infection prevention and control statistics and experts in the field, also confirms the inadequacy in management and enforcement of IPC guidelines [16]. It is known that a positive proactive leadership, support, and presence of senior leaders, team commitment, and clear boundaries of roles and responsibilities are prerequisites for effective action to control of infections in health institutions[16]. For this study, participants highlighted weak management controls on IPC that resulted in failure to implement the IPC plans or establish surveillance for certain HAIs and in Zambia. The infection prevention and control committees barely exist in health institution studied because they lacked committed professionals to take a lead in policy implementation. Contracted companies who are not medical trained personnel have contributed to the failure to meet IPC standards, thus acknowledging challenges on orientation in IPC in the absence of the professional experts to monitor and execute this task despite institution displaying posters with guidelines. This limitation could be addressed by ensuring that the supervisors of the contracted cleaning company’s staff be trained on IPC to ensure adherence to protocols.

## LIMITATIONS

Although results of the study may be used as a learning resource, the study examined issues from the participants’ perceptions and there is an obvious need to complement and extend the work presented with large scale quantitative and mixed-method investigations. Large scale studies will provide data at national level with statistical significance to inform policy on IPC.

## CONCLUSION

Participants for this study demonstrated that the fundamentals of infection prevention standard precautions that included hand hygiene, medical waste management, and bed spacing were not well implemented. While the infection prevention and control committee existed, there was lack of management directive to support monitoring and implementation of IPC policies in the selected sites. Infection prevention and control in healthcare facilities like UTH can be achieved through properly functioning infection prevention and control programs and teams, effective hygiene practices and precautions, including hand hygiene, along with clean, well-functioning environments and equipment.

## Data Availability

All data produced in the present work are contained in the manuscript

## Abbreviations

HAI: Health care Associated Infection
HCP: Health Care Providers
HAIs: Hospital Acquired Infections
IPC: Infection Prevention and Control
IPCP: Infection Prevention & Control Practices
PPE: Personal Protective Equipment
UTH: University Teaching Hospital

## Availability of data and materials

The transcripts generated from the research study are not publicly available due to the sensitive nature of the topic.

## Author’s contribution

MZ, designed and carried out the research, analyzed and interpreted the results and drafted the manuscript. DOC, supervised the student and also contributed in information and application of the critical concepts on Infection Prevention and Control. She also edited the final manuscript. CM, assisted in designing the literature to inform the design and edited the proposal in the early stages. ANH guided the development of the research design, provided guidance on writing the manuscript and read the final version for submission to the journal.

## Acknowledgements

The corresponding author would like to thank the University Teaching Hospital Senior Medical Superintended for access to use the institution to conduct the research. Many thanks to all study participants for their time to take part in the study, Tulani Matenga and Malizani Chavula who assisted with the qualitative software skills for this project and manage the data.

## Ethics approval and consent to participate

The study was approved by the University of Zambia Biomedical Ethics Committee (UNZABREC) **(*REF. No. 059-06-17*)** and the National Health Research Authority (NHRA) in Zambia ***(MH/ 059-06-17)***. Signed informed consent was obtained from individual participants before each interview.

## Consent for publication

Not applicable.

